# A Qualitative Study on Food Taboos Among Rural Pregnant Women in Bangladesh: Motivators for Adherence and Influencers of Taboo-Breaking Behavior

**DOI:** 10.1101/2024.09.09.24313362

**Authors:** Shahrin Emdad Rayna, Fahmida Afroz Khan, Sharraf Samin, Saiqa Siraj, Saika Nizam, Syed Shariful Islam, Md. Khalequzzaman

## Abstract

Understanding the influence of cultural practices on maternal health is crucial in addressing the nutritional challenges faced by pregnant women in rural Bangladesh. Despite significant improvements in maternal and child health indicators, food taboos remain prevalent, impacting the nutritional well-being and health outcomes of vulnerable populations. This study explored food taboos and factors related to their adherence or violation, among rural pregnant women in four districts of Bangladesh: Habiganj, Bhola, Rajshahi, and Cumilla. A qualitative cross-sectional study was conducted with 90 participants through 29 in-depth interviews and 11 focus group discussions. Participants included 21 pregnant women, 23 mothers-in-law, 20 husbands, and 26 healthcare workers. The data were thematically coded and the narrative was analyzed. All participants identified at least one food item restricted by family elders, often based on beliefs about the negative effects of certain foods on pregnancy and the baby’s health. Commonly restricted animal source foods included white carp, trout, duck meat, and mutton, due to fears of convulsions, speech disorders, or undesirable traits in the baby. Raw papayas and pineapples were avoided due to beliefs they could cause miscarriage. Adherence to these taboos was related to the pregnant mother’s preference for vaginal delivery, desire to avoid harm to her child, and profound respect for her elders. Factors enabling the breaking of food taboos included nutritional counseling by health care workers, increased family understanding of maternal nutrition, reduced reinforcement of taboos, and the lack of negative consequences from consuming tabooed foods. The findings highlight the importance of utilizing scientific evidence to challenge food taboos by developing new strategies or updating community-based nutritional counseling programs. Furthermore, including family members and community elders in these efforts is crucial for creating a conducive environment that facilitates dietary changes for pregnant women.

## Introduction

Existing food taboos in the community are particularly associated with women and their special events in life, such as menarche, marriage, pregnancy, childbirth, lactation, et cetera [1]. In most communities’ food restrictions are generally imposed during childbirth and early lactation period of a woman, which usually do not comply with the modern biomedical concepts regarding the proper types and amount of food that pregnant women need to assure optimal maternal nutrition [2]. This ultimately results in suboptimal dietary practices of the pregnant, leading to a state of biological competition between the mother and the fetus in which the well-being of both is at serious risk [3]. Maternal undernutrition is a major public health and development concern in Bangladesh because more than half of the pregnant women in rural Bangladesh, had reduced dietary intake during their pregnancy [4] and the identified reasons behind them were, presence of local beliefs regarding food taboos during pregnancy which was deeply entrenched in the culture of the community, widely practiced, and often their importance were reinforced by the mothers-in-law and elders of the family [4]. Mentally and physically abnormal child, different kinds of illness of the child, and threat of abortion were among the primary reasons for misbeliefs about the tabooed foods [5]. Even though food taboo and their practices are rampant in Bangladesh, very few papers had documented these taboos and the motivations behind them. Using qualitative data from the rural regions of Bangladesh, this study has compiled a comprehensive list of various food taboos practiced, along with the motivators for adherence to the taboos, and a few crucial stimulators in breaking the food taboos.

## Methods

### Programmatic context

The results presented in this paper are part of a formative research initiative taken up by Nutritional International, aimed at informing the development of behavior change interventions to improve the nutritional status of pregnant women in selected districts of Bangladesh. This research contributes to the strategic planning of behavior change programs by identifying key factors influencing dietary behaviors, habits and food taboos, in rural communities. Insights gathered from this study was utilized to create a comprehensive Behavior Change Intervention (BCI) strategy that promotes positive nutritional behaviors among pregnant women. By understanding the motivations behind adherence to food taboos and identifying the influencers of taboo-breaking behavior, this study lays the groundwork for developing culturally sensitive, community-based interventions to improve maternal nutrition and health outcomes in Bangladesh.

### Study type, population and sampling techniques

The initial part of the formative research was an exploratory cross-sectional study using qualitative methods, conducted in four selected districts of Bangladesh named Habiganj, Bhola, Rajshahi and Cumilla (sites where Nutrition International had existing programs). From each of the district, a random upazila (sub-district) was chosen for participant selection. The study population consisted of pregnant women, husbands, mothers-in-law and health care workers (HCWs) operating at the community level. Recruitment for this study began from June 04, 2017 and concluded in March 15, 2018. The pregnant women were identified from the registers of all enlisted pregnant women kept by the community clinics (lowest tier of basic health service facilities in Bangladesh) and enrolled for in-depth interviews (IDIs). Husbands and mothers-in-law of pregnant women who were not the family members of the participating pregnant women, were invited to participate in focus group discussions (FGDs). HCWs who were responsible for providing care for pregnant women (health assistants [HA], family welfare visitors [FWVs] and family planning inspectors [FPIs]), were identified and invited to participate, based on their availability.

A total of 109 participants were selected purposively to represent the four upazilas. The maximum variation technique was employed for the selection of the participants, to ensure the diversity of the study subjects with regards to age, socio-economic status, pregnancy status, gestational age and sex [2].

### Data collection procedure

Data were collected using IDIs with pregnant women and HCWs to explore the existing food taboos and understand their reasoning for adhering to it or breaking it. FGDs were conducted separately among groups of husbands and mothers-in-law, to understand the factors which influenced the pregnant women to accept the taboos, and how they were being motivated to break free from the taboos to ensure optimal maternal nutritional practices. In each FGD, 4-6 participants were included which is considered to be an ideal sample size for such qualitative interviews [6]. Sex-specific groupings were done to remove any male influence over the discussions. An interview guideline was developed to conduct the interviews in a systematic and semi-structured manner. It helped the interviewers to guide the conversations around the existing dietary taboos during pregnancy, and the dynamics of the individual, societal and/or environmental factors influencing it. The qualitative guideline was initially pre-tested in an upazila, close to Dhaka but with similar socio-demographic profiles of our study areas. Based on the findings and consultation with the research team, the questions were modified to reflect literacy levels and cultural interpretations. The IDIs and FGDs were conducted in Bangla by the field team following an interview guideline. Field notes were taken by one of the research assistants, to note down the observations of the surrounding environment while conducting the interview, and any non-verbal reactions to the questions/discussions, which were used in analysis later. To minimize the possibilities for misinterpretation of the participant narratives, extensive discussions with the research assistants and field supervisors were held at the end of data collection to compare notes, discuss findings and to clarify local language and meanings where necessary.

### Data analysis

All the interviews were audio recorded, which was followed by verbatim transcriptions of the interviews, and then translated into English. In order to ensure the consistency of the translation with the original, the researchers double checked the audio with the verbatim and translated materials, to avoid any errors. Thematic analysis was utilized to manually analyze the textual data (i.e.—no textual software was used e.g. Atlas-ti or Nvivo) [7].Open codes were generated, which was followed by coding of the data by reading and re-reading all the transcripts. After completing the initial coding of the interviews, codes were collated under potential themes. Some sections of the interviews were quoted verbatim, and some were modified to enhance readability. The validity and refinement of the themes and its contents were performed after reaching a consensus with the authors and the research team.

### Ethical considerations

This study was conducted according to the guidelines laid down in the Declaration of Helsinki and all procedures involving research study participants were approved by the Institutional Review Board of Bangabandhu Sheikh Mujib Medical University, Dhaka, Bangladesh (BSMMU/2017/3281). All participants provided written informed consent before the interviews began. Utmost care was taken to ensure the comfort of the pregnant women and their children by the data collectors. Sensitive questions or issues were dealt very carefully without hurting the sentiments of the participant and their family. All the members of the data collection team and analysis team cautiously maintained the confidentiality and anonymity of the participants throughout the study.

## Results

A total number of 29 IDIs and 11 FGDs were conducted (Table 1). Among them, there were 21 pregnant women, 23 mothers-in-law, 20 husbands and 26 HCWs.

**Table 1:** Interview methods used and sample size.

| Methods | Type of Participants | Number of<br>interviews | Number of<br>participants |
| --- | --- | --- | --- |
| In-depth interviews | Pregnant women | 21 | 21 |
|  | Health care workers | 8 | 8 |
| Focus group discussion | Mothers-in-law | 4 | 23 |
|  | Husbands | 4 | 20 |
|  | Health care workers | 3 | 18 |

The mean age of the pregnant women who participated in this study was 24.35 ±5.0 years, and their mean age during marriage was 16.48 ±2.3 years. Majority of the pregnant women (61.9%) had completed only until primary education and were homemakers (85.7%).

**Table 2:**
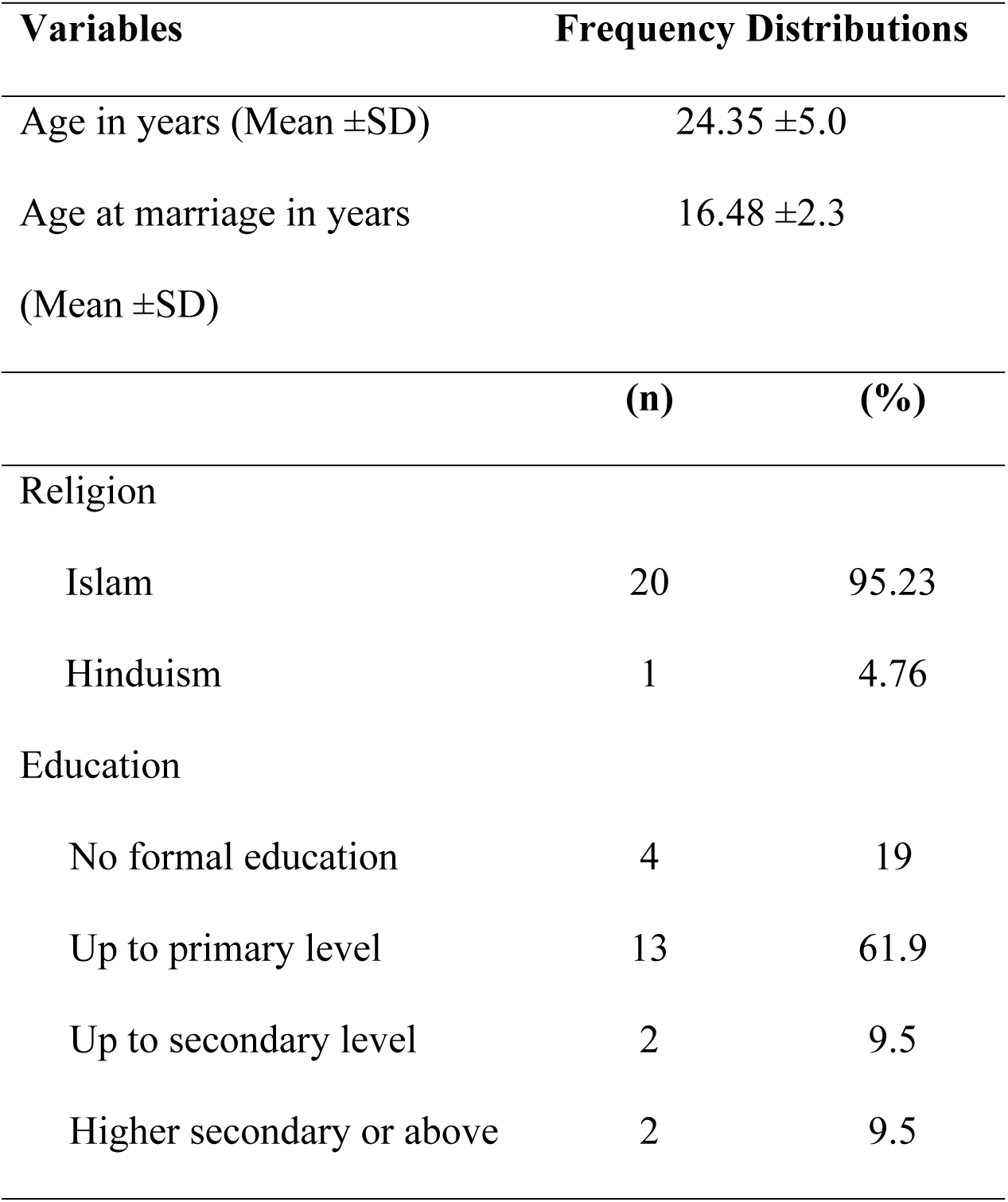
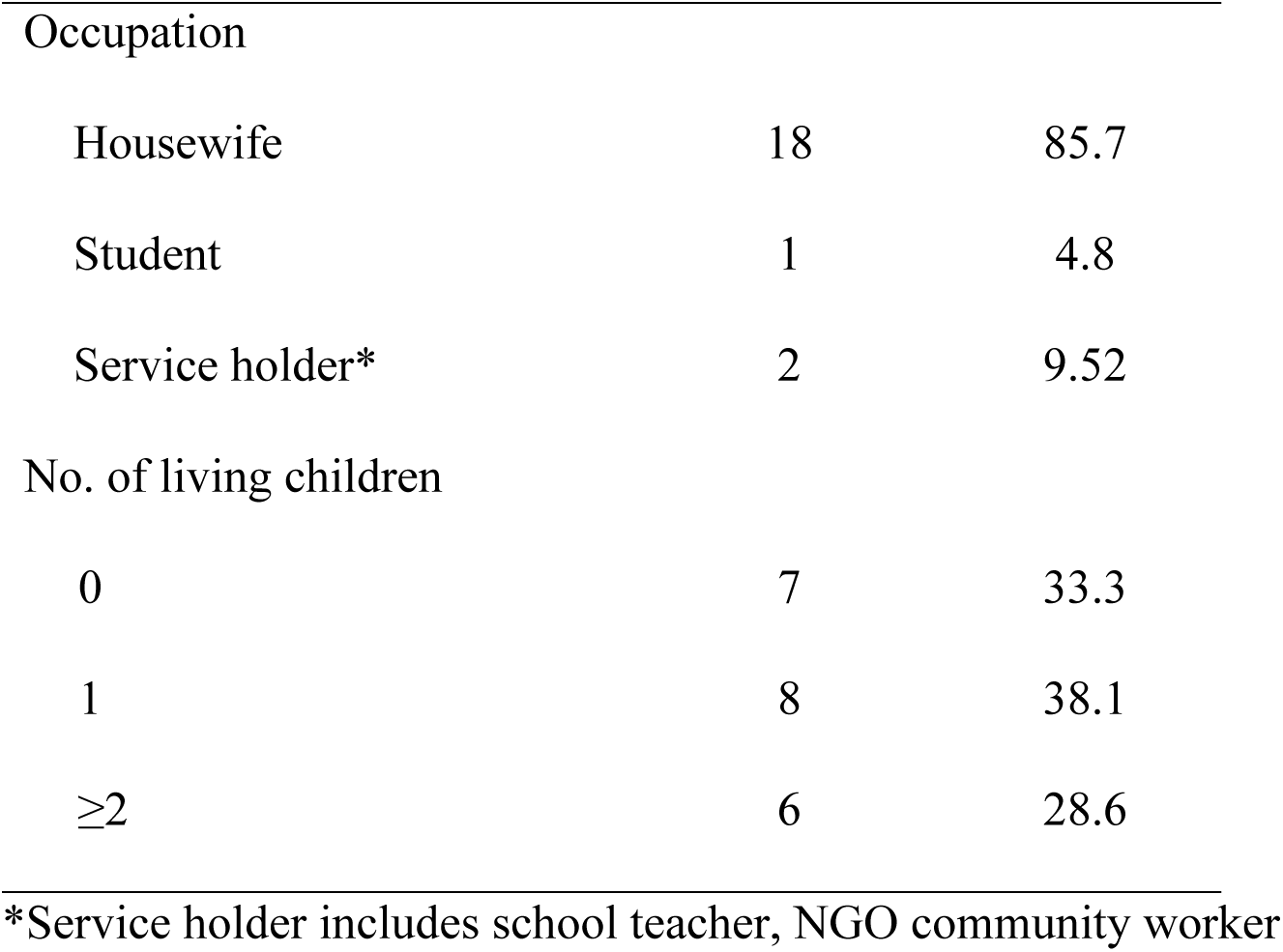
Demographic profile of the pregnant women (n=21)

The various themes which emerged from the interviews and discussions are presented and discussed below.

### Food taboos during pregnancy

Food taboos practiced by the rural women were classified into three groups: animal source foods, vegetables and fruits, and other items. All the participants mentioned at least one food item which was not allowed by her elders or other family members, and many of them modified their diet or eating habits at various levels during pregnancy based on the food taboos.

#### Animal source foods

The current study revealed that majority of the women were prohibited to eat varieties of animal source foods, among which different types of fishes were the most mentioned. The frequently voiced fish was the white carp (locally called in Bangla *Mrigel maach*), which was avoided by almost all pregnant women because it was believed to cause convulsion/ epilepsy of the child (*mrigi rog*).

> “…all the pregnant women in this community avoid white carp (*Mrigel maach*), it is said that this fish causes convulsions of the child…who wants to see that happen to their child? No one!” – IDI, pregnant woman, Cumilla

Additionally consumption of trout fish, yellowtail catfish were also restricted as they were believed to cause convulsions of the baby. The participants from Cumilla mentioned avoiding the zigzag eel (*Baim maach),* as they believed it caused increased fetal movements in their womb; i.e., the baby keeps rolling and twisting in their tummy leading to discomfort of the mother. Similarly, participants from Bhola mentioned avoiding white carp, trout fish, yellowtail catfish for the obvious reasons, but they also included swamp barb (*Puti maach*) and wallago attu (*Boal maach*) in their list. Intake of the swamp barb was thought to cause insufficient breast milk production after delivery, whereas eating wallago attu would cause the baby’s mouth to resemble like that of the fish.

Among other animal source foods, duck meat and mutton were specifically restricted for women living in Cumilla and Bhola regions. Duck meat was avoided as it was believed to cause speech disorders (stuttering) or changes in the voice quality (harsh voice like duck) of the baby. Likewise, eating mutton during pregnancy might result in the baby having a bad smell or body odour (*botka gondho*).

> “…my mother-in-law told me not to feed duck meat… apparently if you consume duck meat then the baby’s voice breaks, the voice is not of good quality” - FGD, mothers-in-law, Cumilla

> “… some women don’t want to eat mutton, and when I asked them why… they say that the meat has a stench, and it is believed that when the child grows up, also smells foul.” – IDI, HCW, Cumilla

#### Fruits and Vegetables

Fruits and vegetables were the most consumed food items by all the pregnant women because it was inexpensive and widely available. Yet, raw papayas and pineapples were named as restricted food items because they were reported to cause miscarriage. Some respondents also mentioned that consuming raw papayas will result the baby’s skin to crack (*fata*), and if pineapple is consumed it will turn the baby’s hair red.

> “…one shouldn’t feed pregnant women raw papayas… some say you lose the baby…” –FGD, mothers-in-law, Bhola

> “raw papayas we always eat it cooked…if not child is born cracked… once my daughter-in-law ate raw papayas in front of me, I was very afraid for her then…” – FGD, mothers-in-law, Rajshahi

Other fruits and vegetables which were not preferred for consumption were black eggplant, indian spinach, coconut and butternut squash as they possibly caused allergy, cold, asthma (*hapani*) in the newborns. A few participants also mentioned that drinking coconut water was not allowed as it can turn the baby’s eye color to red and/or opaque (*chok laal/ghola hoye jae*).

#### Other food items

Rice and its variations remain the staple food in all the meals consumed by the pregnant women. Yet, the intake of a few forms of rice were restricted during pregnancy. A few participants mentioned avoiding eating cooked rice soaked in water overnight (*panta bhaat*), as it was considered as a cold food which might lead the newborn catching a cold. Also, eating uncooked rice, puffed rice (*muri* and *khoi*) was not allowed as it is believed to cause insufficient breast milk production later on. Expecting mothers from Rajshahi also avoided eating puffed rice, rice crisps (*chira*) and fried rice powder (*chaatu*), because the elders believed, these foods when consumed will coat the body of the fetus with a very sticky greasy material which will be very hard to clean after the baby is born.

Consuming any cold food or drinks was barred for the pregnant women of Cumilla and Rajshahi as they believed cold foods will cause common cold of the baby. Additionally, women from Cumilla mentioned that drinking cold water just before delivery makes the delivery difficult as the baby gets stuck in the womb and cannot come out via normal vaginal delivery. In such cases the mother needs an episiotomy/caesarean section to deliver the baby, which was not favored.

> “While I was pregnant, people told me to not eat cold foods… since I have asthma, I was afraid that drinking or eating cold foods will cause my child to have a cold…or worse… pneumonia…” - IDI, pregnant woman, Rajshahi

### Motivators for adhering with food taboos

Many participants in this current study did realize that many reasons behind avoiding most of the tabooed foods were probably insignificant and had no scientific validity, but they still followed them out of fear for their perceived negative effects. Few of the reasons identified for adhering with the taboos included: preference for vaginal delivery for childbirth, to avoid the financial burden of a caesarean section, to avoid causing any harm to their baby, and deep trust on their elders.

The preference for a normal vaginal delivery either at home or an institution was mentioned by majority of the participants. Despite knowing the importance of nutritious food for the baby’s well-being, many participants avoided eating too much, fearing a large baby would lead to delivery complications and the need for a costly caesarean section. Conversely, some families encouraged more food intake, believing that a fuller stomach would limit space for the fetus to grow, making normal delivery easier and reducing the need for a caesarean section.

> “The in-laws tell them if they eat more the baby will be bigger and it will take a cesarean section to deliver the baby, which they cannot afford to do. It is one of the major reasons for eating less…” – IDI, HCW, Rajshahi

> “I try to eat more now, so that my belly remains full… everybody suggests me to eat more food so that the baby will remain small but also healthy… and I will not need caesarean section” – IDI, pregnant woman, Cumilla

Additionally, the participants wanted to avoid caesarean section or hospital delivery was due to the financial burden it would impose on the family, and the extended time it would take for a mother to recover also. Similar sentiments were also echoed by the husbands, where they explained, because of their meager earnings they could not afford caesarean birthing and hence, the expecting mothers complied with eating less amounts of food when told to. Additionally, another husband mentioned that, undergoing caesarean delivery might be a matter of embarrassment for the new mother, as during the postpartum period she is unable to contribute much to her household chores or family income.

> “The mother is asked to eat less because, when the child gains more weight, it creates problems during the delivery… also, the woman usually works to earn for herself and her family. When the baby is born by cesarean birth, the mother cannot earn or do her daily chores like before. She then feels ashamed and keeps her head bowed down in front of the family.” – FGD, husbands, Habiganj

Another key reason for adhering to the taboos was the unwavering trust many participants placed in their family elders, particularly the female heads of the household, who were regarded as more knowledgeable and experienced due to their own experiences with childbirth.

> “My mother-in-law gives me food that is good for me, and doesn’t give those food that aren’t good for me… I don’t say anything else” – IDI, pregnant woman, Habiganj

Many HCWs also considered the blind faith the daughters-in-law had on their mothers or mothers-in-law as the prime reason for following the taboos.

> “… still now in villages, the daughters-in-law have blind faith on their mothers-in-law… it’s not blind faith …it’s like compulsion, despite of our counselling, they (pregnant women) say, I will do whatever my mother-in-law asked me to do!” – FGD, HCWs, Bhola

### Influencers for breaking the food taboos

Even though food taboos are widely prevalent in rural communities, the interviews revealed that many pregnant mothers did not follow this cultural practice because they were not reinforced by their family members or community elders.

> “I ate mutton several times during my pregnancy… many people told me that if I eat mutton a bad smell will come from my baby’s body after birth. But I ate it when I was pregnant with my eldest child, and nothing was wrong with my child after his birth…” - IDI, pregnant woman, Cumilla

Some mothers also pointed out that one of the reasons for breaking food taboos was that the baby or mother did not experience any of the adverse effects of the particular tabooed food, even after consuming them.

> “During my first pregnancy, everyone forbade me to eat too much food…they used to say that if I eat too much food, the baby will get bigger and I will face difficulties during delivery…but I didn’t listen to them, I ate as much as I wished and delivered a healthy baby per vaginally” – IDI, pregnant woman, Rajshahi.

Many participants also highlighted the lack of enforcement to the adherence of taboos. This was because of the counselling by the HCWs to dispel many long standing food taboos, improved knowledge of the mother-in-law and husband regarding maternal nutritional, and raised educational level of the rural people. It was reported that:

> “In our days we (elderly women) were only given bitter cumin paste or coriander paste to eat after our deliveries… it was meant to purify our bodies after the delivery of the baby …then we had no idea about what to eat and what not to… now days the doctors *(field HCWs are called as doctors)* advise us. The old days are gone. Now we feed the expecting mother ducks, eggs, fish…” - FGD, mothers-in-law, Cumilla.

> “Before the pregnant women were given rice husks (locally called as *chaff*) to eat… it was the rules of the society… the old people did not understand many things then… but now everybody is more aware… everybody wants the mother and the baby to be healthy… that’s why we listen and follow what the health worker tells us…” - FGD, husbands, Cumilla.

Nutrition focused counselling are usually provided to pregnant mothers by HCWs during their ante-natal visits at the health center, which is also supplemented with door-to-door household visits by domiciliary workers and by conducting yard meetings (*uthan boithok*). During these sessions, the mothers and their family members are informed about what to eat, how much to eat, and the impact it has on the mother’s and child’s health. These are aimed not only for improving the awareness regarding the nutritional needs of the mother, but the HCWs also aspire on abolishing the long standing food taboos in each region.

> “…as we are providing healthcare related information, the bad culture of adhering to food taboos is mostly gone. Like before when they used to refrain their daughters-in-law against eating too much, but now because of us advising them they are providing proper food and care to their daughters-in-law.” – IDI, HCW, Bhola

The ability of the rural people to accept this new information which contradicts their preexisting beliefs on food taboos is influenced by the improved educational status of the expectant mother and her husband. Likewise, hearing and viewing health messages on the radio and television, as well as observing other people in similar situations.

> “Before there were many misconceptions about food… but now they are being removed… because either the wife or the husband is educated… so when we tell them something, they understand it. The husband then shares the new knowledge with his parents, helps them to change their views. The times have changed … the parents also listen to their kids now.” – IDI, HCW, Cumilla

> “People said eating barb fish (*Puti maach*) and duck meat hinders production of breast milk after delivery. But I don’t believe these and didn’t follow them… you see, I am a school teacher… I teach students about nutritious food from the school’s science book. I also suggest my neighbors to eat food like me. I tell them, those are nutritious food, and you should eat this food every day.” – IDI, pregnant woman, Habiganj

## Discussion

This study aimed at exploring the existing food taboos in the rural areas of Bangladesh among the pregnant women, along with the influencers for either adhering or breaking the taboos. The results revealed that all pregnant participants were at least familiar with one food item which was tabooed during pregnancy. These beliefs are an integral part of the oral traditions of Bangladesh’s rural communities, where elders pass on indigenous knowledge and beliefs from generation to generation [8]. Therefore, women already have developed detailed perceptions and belief systems related to pregnancy and childbirth, prior to antenatal care and nutrition education contact [4]. The current study findings indicate that various food items from animal source were most commonly avoided. Fishes like white carp, trout fish, zigzag eel, were not consumed in rural Bangladesh, as they were believed to cause increased movement of child/ convulsions and fetal malformation. Correspondingly, in countries like Tanzania [9], Indonesia [10], and Malaysia [11], fishes like eel, catfish, shrimp, stingray, mackerel, shellfish, were tabooed because they were believed to cause difficulties during delivery. Meat like duck meat and mutton, were avoided by the participants of this present study as they were perceived to cause respiratory disorders of the baby. The consumption of meat, chicken and eggs during pregnancy was also found to be restricted in Indonesia [10], Nigeria [12,13] and South Africa [14], because the mothers believed the child maybe born disabled and develop bad habits. Other than protein foods, the most frequently tabooed food items identified in this study were pineapples and green papaya, which was perceived to cause miscarriage. These fruits were not only avoided in Bangladesh, but also in various parts of India and other Asian countries due to the fear of abortion [1,5,15,16]. On the other hand, in African countries, women cited oranges, mangoes, peaches, and other yellow/orange colored fruits as tabooed, as they believed that the baby would then be born jaundiced, a condition causing the newborn’s skin and eyes to become yellow [14,17]. Most of the taboo foods reported in this study are rich sources of essential nutrients and micronutrients, which are crucial for maternal health and child development. Restricting these foods from the pregnants’ food basket, limits their dietary diversity and leaves them vulnerable to many nutritional deficiencies during pregnancy, resulting in the potential detriment of both mother and the unborn child [14].

The participants’ reasons for adhering to food taboos in this study were, not wanting to harm their baby, preference for vaginal delivery as the birthing process, and blind faith on their elders. These reasons were also cited in previous studies done in Bangladesh [4,5,18] and India (West Bengal) [1]. The pregnants also mentioned that, although some of the reasons for food taboos were trivial and sometimes based on mere speculations, they still uphold them for fear of their perceived repercussions. Similar reasons for adhering with food taboos was also reported by Arzoaquoi et al. (2015) [2] and Haslam, Lawrence, & Haefeli (2003) [19], where the participants were aware of the reasons for adhering to the taboos, and such practice was observed as positive antenatal health behaviors within the community favoring positive pregnancy outcomes. Among other reasons quoted were, a symbol of respect for elders [2,17], mother’s own previous bad experience or of others [10], maintaining harmony with entities, natural and supernatural [20], preventing any misfortunes or calamity from happening [19], avoid being excommunicated from their own family or community, and sustaining a feeling of belonging or cohesion with their community and cultural values [2].

Despite of the prevalence of food taboos in the rural communities in Bangladesh, this study was able to button down participants who did not follow any food taboos. A notable reason behind breaking this cultural practice was the improved knowledge of the mothers-in-law and husbands on maternal nutrition, which in turn led to reduced reinforcement of the taboos on the pregnant mother by her family members. Likewise, experiencing no adverse effects upon consuming the tabooed food items, and higher educational level of the rural people, enabled the mothers to break the existing culture of taboos. These findings are one-of-a-kind, because previous studies from Bangladesh only informed the existing food taboos and adherence to them [1,4,5,21]. Only one study reported the lack of adherence to food taboos, because of the increased knowledge of healthy eating during pregnancy and gendered empowerment of women residing in an urban slum of Bangladesh [18]. In rural areas, there are numerous ongoing programs emphasizing on nutrition-sensitivity to improve the nutritional status of pregnant women. Among these are counselling sessions which are important tools to break food taboos, because they equip the individuals with the knowledge about the nutritional value of different foods, dispel myths and misconceptions, and helps to create an enabling environment to support the woman’s changed dietary behavior [22,23]. Additionally, an increase in education levels in rural communities has probably led to the questioning of traditional practices and belief, and to seek out information and advice from healthcare professionals. These in particular have resulted in the reduction of the enforcement of food taboos by the family/community members, and an increase in the uptake of ante-natal care services [24], which can give pregnant women more freedom and assurance to make their own food choices, including food items that were previously considered taboo. The results from this study might be useful while developing strategies to encourage dissent towards the food taboos, by utilizing scientific evidence to dispel cultural or religious prohibitions against the tabooed items. Furthermore, the findings can be integrated in the existing community-based nutritional programs as an effort to create more awareness. In conjunction to these, mechanisms can be placed to routinely identify women observing food taboos, assessing the reasons and provide innovative means to minimize their negative and maximize their positive nutritional effects [24].

Among the limitations of this study, the findings are based on self-reported maternal dietary patterns, thus recall bias and social desirability bias may have been present. Although the intention of this study was to collect and validate data via participant observation, due to resource limitations, this was not feasible. We are convinced that the findings reflect actual practices, given the consistency of the results across the qualitative interviews and the previous studies discussed.

## Conclusions

This study revealed the existence and practice of food taboos during pregnancy, which varied across the different geographical groups in rural Bangladesh. All the participants were familiar with at least one tabooed food item and were motivated to comply with the taboos because of their desire to protect their unborn child, preference for vaginal delivery, and a sense of profound respect for their elders. On the other hand, nutritional counseling, increased nutritional knowledge of the pregnant women and her family members, reduced reinforcement of taboos, and lack of negative consequences from consuming tabooed foods, enabled the mothers to breaking off the food taboos. It is important to note that breaking a cultural or religious practice can be a difficult decision and not everyone may follow this route. However, it is important to provide a preliminary set of recommendations based on the study findings to address public health problems related to adherence to food taboos and promote healthy lifestyles.

## Data Availability

The datasets used and/or analysed during the current study are available from the corresponding author on reasonable request.

## Acknowledgements

The authors would like to acknowledge the support of the team from Nutrition International for all their technical and administrative support. The authors would also like to thank all the participants for their valuable time and assistance in sharing their views for this study.

